# Sequencing Data of North American SARS-CoV-2 Isolates Shows Widespread Complex Variants

**DOI:** 10.1101/2021.01.27.21250648

**Authors:** Colby T. Ford, Rachel Scott, Denis Jacob Machado, Daniel Janies

## Abstract

Several new variants of the SARS-CoV-2 have been isolated in the United States, Mexico, and Canada. Many of the variants contain single variants of functional significance (e.g. S: N501Y increases transmissibility). To study the occurrence and co-circulation of these variants, we have developed an easy-to-use dashboard at janieslab.github.io/sars-cov-2.

We created a multiple sequence alignment workflow and processing script to generate a variant dataset, which populates this dashboard. We then use the features of the dashboard, such as visualization of the single and complex nucleotide variants geospatially and in a color-coded matrix format. Users also interact with the dashboard to filter the underlying data to regions of interest and or variants of interest. The user can export reports based on the desired filters, which we intend to be used for regionally specific pandemic response. We find in Genbank, an isolate from Massachusetts containing [(S: Q677H), (ORF3a: Q57H), (M: A85S), (N: D377Y)] collected on September 11, 2020.

Moreover, we find that many viral isolates bear a marker of increased transmissibility (S: N501Y) in linkage with at least one variant of concern isolated from Ohio also range across the Untied States and stretch from British Columbia, Canada to Mexico. When we analyze co-circulation of more complex variant constellations with (S: N501Y), we note that the Upper Midwest and Northeast United States contain these isolates.

In summary, the viral variants that have raised concern in a few US States in recent reports are widespread. Based on the increase in the proportion of variant viruses being sampled and some empirical evidence in the United Kingdom, South Africa, and Ohio, these variants are likely to lead to increased transmission of SARS-CoV-2 across North America in the coming months.

## Introduction

The United States has experienced the arrival of SARS-CoV-2 variants containing mutations such as Spike N501Y that increase transmission of the virus in the United Kingdom and South Africa (1).

In addition, there have been reports of SARS-CoV-2 variants containing novel mutations in the coronavirus’ Spike protein and other single variants in low frequency locally [e.g. (S: Q677H), (M: A85S), and (N: D377Y)]. This is shown in Table 1A in Tu et al. (2021).

**Table 1.** Sample data format of accession variant information for mapping and visualization.

| Accession / GISAID Entry Name | Mutation | Abbreviation | State | Country | Latitude | Longitude |
| --- | --- | --- | --- | --- | --- | --- |
| hCoV_19_USA_OH_CDC_2_3693476_2020_EPI_ISL_747131_2020_11_04 | 21575: C>T (S: L5F) | OH | Ohio | USA | 40.388783 | -82.764915 |
| hCoV_19_USA_OH_USAFSAM_S241_2020_EPI_ISL_812576_2020_07_27 | 21575: C>T (S: L5F) | OH | Ohio | USA | 40.388783 | -82.764915 |
| MW454623_USA_MA_MGH_03165_2020 | 23593: G>T (S: Q677H) | MA | Massachusetts | USA | 42.230171 | -71.530106 |
| hCoV_19_USA_OH_DEC106_2020_EPI_ISL_826463_2020_12_30 | 23593: G>T (S: Q677H) | OH | Ohio | USA | 40.388783 | -82.764915 |
| MT993885_USA_VA_DCLS_1271_2020 | 25563: G>T (ORF3a: Q57H) | VA | Virginia | USA | 37.769337 | -78.169968 |

The properties of this complex variant containing [(S: Q677H), (M: A85S), and (N: D377Y)] has not yet been studied in empirical tests. However, the authors report that like Spike N501Y-containing variants the United Kingdom and South Africa, [(S: Q677H), (M: A85S), and (N: D377Y)]-containing variants have increased in frequency in Columbus, Ohio from from 10% in December 2020 to upwards of 61% in January 2021. In another preprint by Pater et al. (2021), authors report additional isolation of the variant containing [(S: Q677H) and (N: D377Y)] in Michigan, Utah, and Texas.

## Results

Using our SARS-CoV-2 dashboard, we report on several isolates of interest put in the public domain in NIH’s Genbank (e.g., MW454623.1 by Lemieux et al.) and several isolates in GISAID.

This Massachusetts isolate contains: (S: Q677H), (ORF3a: Q57H), (M: A85S), (N: D377Y), as (like another from Ohio posted in GISAID (EPI_826463) reported in Tu et al. (2021v2). The Massachusetts isolate was collected on September 11, 2020. The Ohio isolates in public databases were collected primarily in December 2020.

Here we extend these works by defining a new constellation of viral mutations in SARS-CoV-2 variant [(S: Q677H), (ORF3a: Q57H), (M: A85S), (N: D377Y)] currently recorded in Massachusetts and Ohio from public databases. See Figure 2.

### Additional Observations

(See Figures 1 and 3.)

- We find [(ORF3a: Q57H), (S: Q677H)] isolates in Massachusetts, Michigan, Minnesota, and Ohio.
- Isolates of [(ORF3a: Q57H), (S: Q677H), and (N: D377Y)] occur in Michigan, Minnesota, Utah, and Wisconsin.
- We find [(S: N501Y), (ORf8: R52I)] isolates widespread in the United States and with records for Canada and Mexico, including: British Columbia, California, Connecticut, Florida, Illinois, Maryland, Wisconsin, Minnesota, New Mexico, New York, Oklahoma, Ontario, Oregon, Pennsylvania, Quebec, Tamaulipas Mexico, and Texas.
- We find [(S: N501Y), (N: D377Y)] isolates also widely distributed including: British Columbia, California, Colorado, Connecticut, Florida, Georgia, Illinois, Maryland, Michigan, Minnesota, New Mexico, Ohio, Oklahoma, Ontario, Oregon, Quebec, and Texas.

**Fig. 1.**
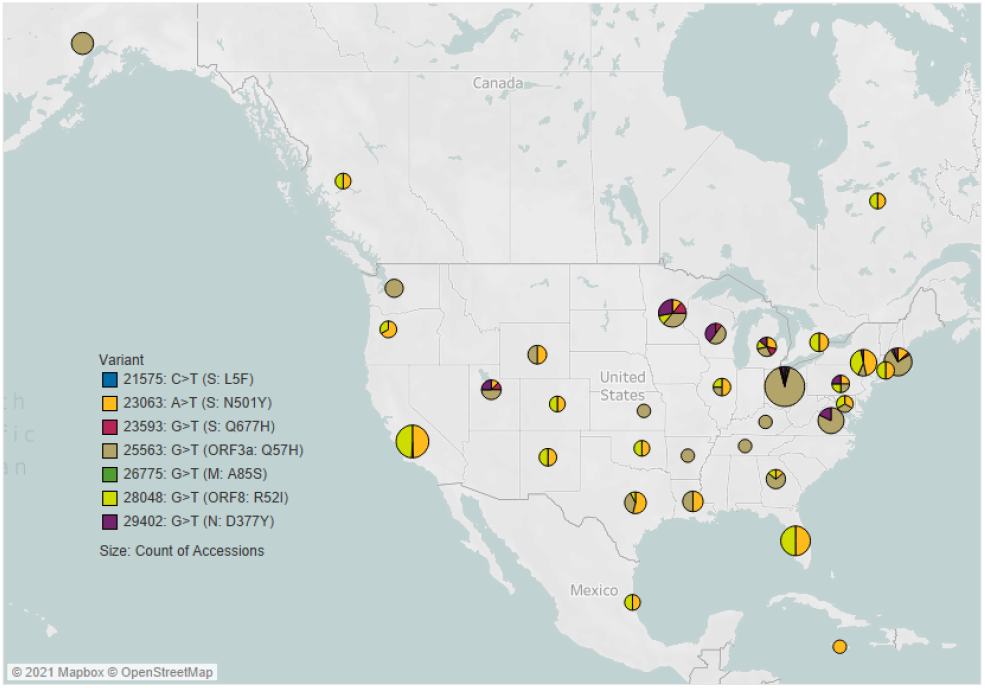
Map breakdown of SARS-CoV-2 variants. **Note:** Tu et al. (2021), in Table 1A, presents a minor single nucleotide variant that they list as A25563G. However, in the data we have, this position contains the major SNV: G25563T. This SNV is very common as it occurs in hundreds of isolates from North America. SNV G25563T corresponds to amino acid substitution Q57H in ORF3a and has risen in preponderance in Europe and the USA over 2020 (4). As such, we have corrected this variant as G25563T, Q57H in ORF3a in our analyses.

**Fig. 2.**
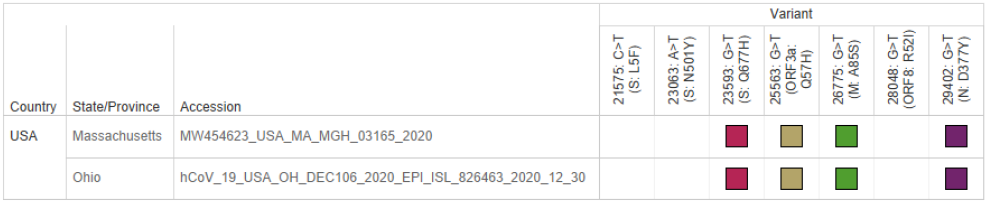
Unique variant constellation currently only recorded in Ohio and Massachusett

**Fig. 3.**
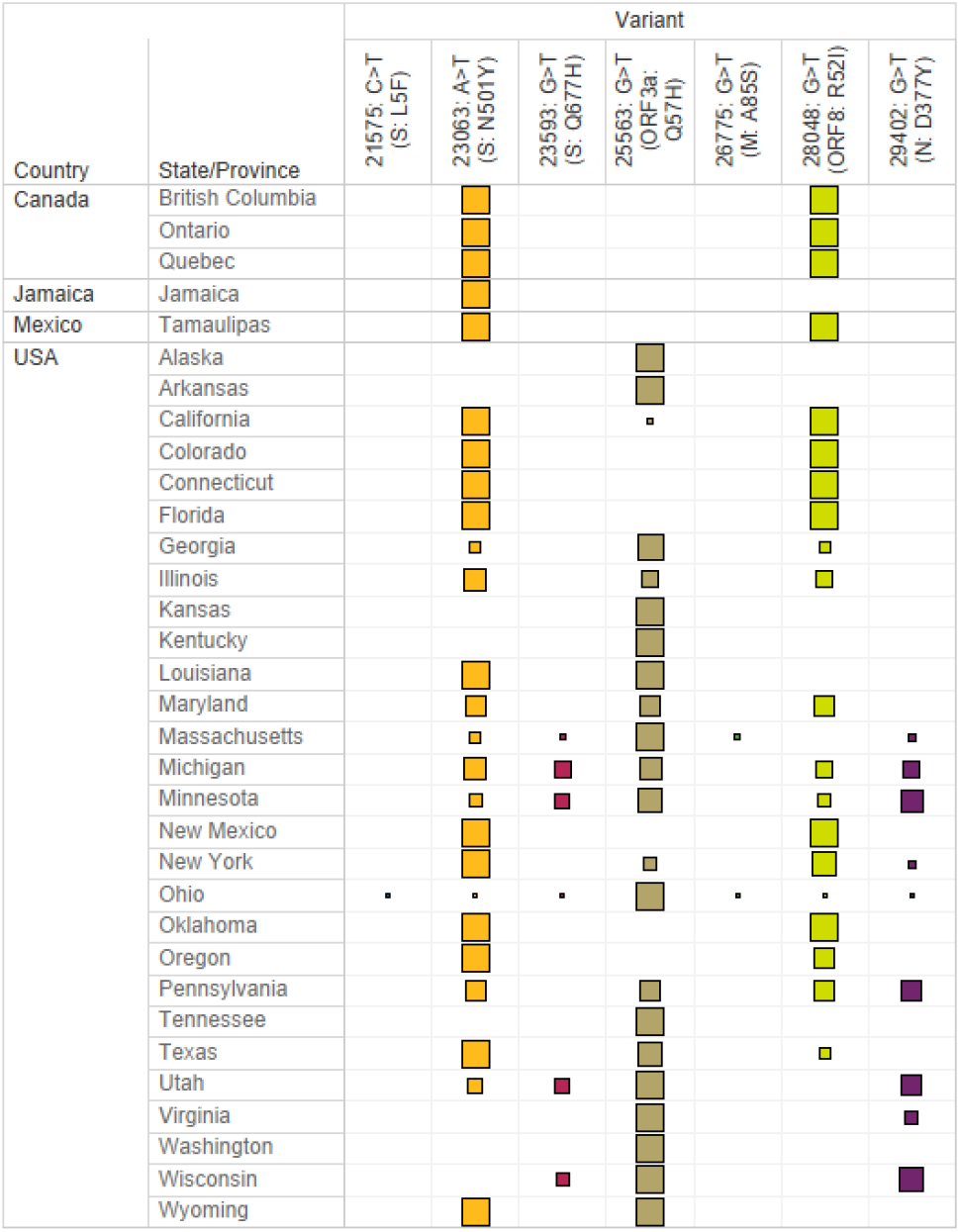
Color-coded matrix format table illustrating of complex variant occurrences by location. (Point size represents percentage of isolates with each mutation. This is a summary, please see dashboard for details.)

We expect these lists of places containing these mutational constellations to continue to increase in content and geography, and will update the SARS-CoV-2 dashboard.

## Conclusions

Here we show that the variants that contain mutations associated with increased transmissibility such as N501Y of the UK and South African variant occur widely in North America. [(S: N501Y) linked to (ORF8: R52I)] or [(S: N501Y) linked to (N: D377Y)] in viral isolates is much more geographically widespread than initially reported.

These N501Y-bearing isolates are in co-circulation and there are several other variants that such as [(S: Q677H), (ORF3a: Q57H), (M: A85S), (N: D377Y)], [(ORF3a: Q57H), (S:Q677H)], [(ORF3a: Q57H), (S: Q677H), and (N: D377Y)] in the states of Massachusetts, Michigan, Minnesota, and Ohio. As such, these viruses can recombine and or experience selective pressure that leads to more complex variant constellations with functional consequences.

## Methods

We downloaded data from the GenBank (5) and GISAID (6, 7) public repositories with dates of isolation ranging from March 2020 to early January 2021 with the exception of the reference isolate from Wuhan in December 2020. We aligned the data using MAFFT v7.310 (8) and edited out spurious ‘n’ characters added by data submitters. We used the Wuhan-Hu-1 isolate (NCBI’s GenBank accession number NC_045512) as an ungapped nucleotide reference sequence, counting from 1, to create a single .NEXUS file of 349 taxa and 29,912 characters. We did not curate the alignment beyond the last mutational position of interest, 29,402.

### Bioinformatics Data Preparation and Visualization

Using AliView (9), we selected mutational positions of interest and set each position aside as individual .FASTA files containing just the accession data and one base.

Once .FASTA files of the individual mutations of interest were created, these files were parsed using a custom R script. This script creates a data frame of each accession, position, reference nucleotide, and variant nucleotide and also parses out location information from the accession. We then join these data with a reference table of states, countries, and latitude and longitude information. See Table 1 below for an example.

Tableau was used as the visualization engine and provided an interactive way to plot variant occurrence geospatially and provides an intuitive mechanism to browse accessions by location or variant. The color-coded matrix format is much easier to visualize complex variants than tree-based formats. See Figure 4.

**Fig. 4.**
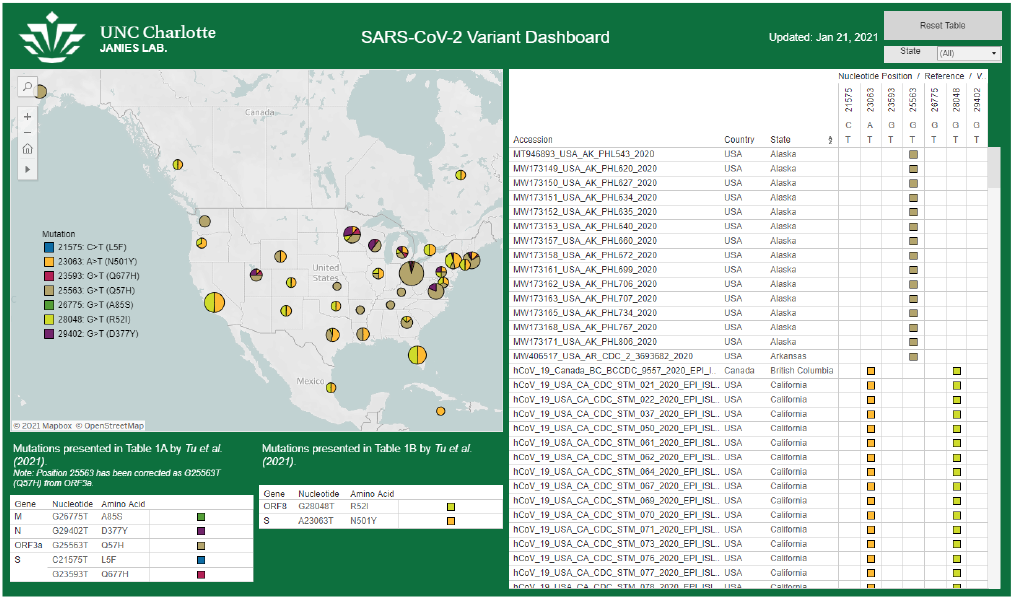
Screenshot of the SARS-CoV-2 Variant Dashboard

## Data Availability

All code, current data, and acknowledgments are available on GitHub at https://github.com/janieslab/SARS-CoV-2_VariantDashboard.
To view the interactive dashboard of current variants, visit https://janieslab.github.io/sars-cov-2.

https://janieslab.github.io/sars-cov-2

## Data and Code Availability

All code, current data, and acknowledgments are available on GitHub at github.com/janieslab/SARS-CoV-2_VariantDashboard.

To view the interactive dashboard of current variants, visit janieslab.github.io/sars-cov-2.

## ACKNOWLEDGMENTS

We thank all the data submitters and have included acknowledgment lists in the supplemental GitHub repository. We acknowledge support from UNC Charlotte (College of Computing and Informatics, Department of Bioinformatics and Genomics, Bioinformatics Research Center, Research and Economic Development, Government Relations, University Research Computing, and the School of Data Science). The views expressed here reflect only the authors’ views and not necessarily the views of those acknowledged.

## Notes

### Competing Interest Statement

The authors have declared no competing interest.

